# Multitask learning from clinical text and acute physiological conditions differentially improve the prediction of mortality and diagnosis at the ICU

**DOI:** 10.1101/2020.06.30.20143677

**Authors:** L.G. Reichmann, G. Valdes, Romain Pirrachio, Y. Interian

## Abstract

The prediction of mortality of critically ill patients has stimulated the development of many severity scoring algorithms. The majority of the models use physiological measurements obtained during the first hours of admission (i.e., heart rate, arterial blood pressure, or respiratory rate). In this study, we propose to improve the performance of current scoring system by including free text from patient’s medical history. Although the primary outcome was in-hospital mortality, we chose a model architecture to provide simultaneous assessment of ICD-9 codes and groupings. We hypothesized that including patients’ medical history with a multitask learning approach would improve model performance. We compared the predictive performance obtained with our approach to the best models previously proposed in the literature (baseline models). We used the MIMIC publicly available database which includes > 60,000 ICU admissions between 2001 and 2012. The patients’ condition at admission was accounted for by the preliminary diagnosis at admission and the medical history extracted from the discharge summaries notes. Unstructured data was processed through a Gated Recurrent Units layer with pre-trained word embeddings, and the hidden states were concatenated to the remaining structured-tabular data. Baseline models achieved similar results than in previously published work, but our artificial neural networks models showed significant improvement towards classification of mortality (AUC-ROC = 0.90). Including the medical history improved all tasks but relatively more the ICD-9 codes prediction than the mortality. The clinical prediction model presented here could be used to identify patients’ risk groups, which would improve the quality of ICU care, and further help to efficiently allocate hospital resources.

## 1) Introduction

Efforts to model the prognosis of patients in intensive care units (ICUs) date back to the early 1980s (Le Gall, Lemeshow, and Saulnier 1993, Knaus et al. 1981). Since the 1980s, there has been a significant increase in the availability of electronic health records and open access medical databases. Both of these tools have helped improve the performance of risk scores predicting mortality, length of stay, and/or post discharge outcomes, among others (Johnson et al. 2017).

In most of the cases, scores are calculated from data collected within the first 24h after the ICU admission (i.e., APACHE, SAPS, and MPM). Although there are pros and cons on including more variables, in general one wants simple models to facilitate their wide spread adoption. In fact, one of the challenges of digital medicine is selecting the potential predictors from a rapidly increasing pool of variables, especially since clinical data is being archived at the bedside.

Traditional scoring systems are associated with a number of limitations. First, the selection of score variables and their weights initially relied on subjective methods, e.g., a panel of experts that select variables according to their perceived relevance for predicting mortality (e.g., Le Gall, Lemeshow, and Saulnier 1993). This limitation was overcome by using objective variable selection through statistical modelling techniques (Pirracchio et al. 2015, Rajkomar et al. 2018, Gennatas et al. 2020). Second, the traditional models generate a scoring system, which is then transform to a probability of mortality based on an equation that has been calibrated with specific β-coefficients (Sekulic et al. 2015). A major limitation of these scoring systems is that they require periodic recalibration (Lee and Maslove 2017). With improvements in treatments and technologies, the weights of certain predictors in the model may decrease, increase, or remain unchanged with time. For example, one of the most used scores in clinical practice, the SAPS II score, includes 12 physiologic variables, age, type of admission, plus three co-morbidities variables (Le Gall, Lemeshow, and Saulnier 1993). A newer version of the Simplified Acute Physiology Score (SAPS III, Sakr et al. 2008, Metnitz et al. 2005) added three more comorbidities, and a variable that accounts for the reason for ICU admission. Most of these risk scores models were developed using logistic regression, i.e.,a linear model. However, recent developments in machine learning may allow us to overcome the rigidity of the traditional scoring systems (Pirracchio et al. 2015). A key difference with traditional logistic regression methods, is that data adaptive methods such as gradient boosting, random forest or artificial neural networks (aNNs) can effectively model non-linearities in the function if enough data is available. Moreover, aNNs can not only handle large volumes of data, but also process unstructured data such as text notes from health care providers. The information contained in clinical notes can be extracted in several ways. Some variants of recurrent neural networks, i.e., Gated Recurrent Units (GRU), allow to extract only the relevant information to make predictions, and forget the rest (Cho et al. 2014). In this architecture, words not only get assigned representations, or embeddings, based on their meaning, but also based on their sequence. During training, GRUs find parameters that decide whether to store past information and determine the next output. Since the intermediate states are never observed, they also called “latent” or hidden states.

Machine learning models have shown to improve model generalization when they are trained for more than one task simultaneously. With this approach, called multitask learning, we could use extra outputs available to learn more efficiently related tasks with a shared representation (Caruana 1997). When trained simultaneously on multiple related tasks through multitask learning, backpropagation is done in parallel on all the outputs in the multitask learning net. These characteristics allow aNNs to learn new representations of the relevant factors from the data itself, such as tasks relatedness or disjointness. Overall, multitask learning methods have resulted in better model predictions (Argyriou, Evgeniou, and Pontil 2007, Zhang et al. 2014). In the context of patients’ care, lab test results and diagnoses will become available as patients’ health evolve. At discharge, diagnoses are coded following the International Classification of Diseases (ICD9-CM system, CDC 2011). The ICD9-CM codes are a granular source of diagnose descriptions assigned at the end of a patient’s stay, mostly for budgeting and administrative purposes. Codes have different number of digits depending on their level of detail. Diagnosis codes are composed with either 3, 4, or 5 digits where the first 3 correspond to the heading category that may be further subdivided to provide more detail. The ICD9-CM system has ∼ 13,000 codes, and because several codes could be assigned to the same patient, it is a good system to implement a multitask learning method.

Here, we proposed a clinical prediction model based on a aNN (with and without multitask learning), building on the features used for the SAPS II score, with the addition of clinical text corresponding to the medical history. The primary outcome was in-hospital mortality, yet the model architecture also provides a simultaneous assessment of the 100th most frequent ICD9 codes, and the most frequent ICD9 groupings. We compared the predictive performance of the models with the original SAPS2 score as well as a new logistic regression model, an eXtreme Gradient Boosting model (XGBoost, Chen and Guestrin 2016), and a neural network model.

## 2) Methods

### a) Dataset

We used the Multi-parameter Intelligent Monitoring in Intensive Care (MIMIC) publicly available database, which has been developed by the MIT Lab. The database comprises de-identified detailed clinical information regarding >60 000 stays in ICUs at the Beth Israel Deaconess Medical Center in Boston, Massachusetts, collected as part of routine clinical care between 2001 and 2012. The project’s Institutional Review Board (IRB) was approved by the Beth Israel Deaconess Medical Center (Boston, MA) and the Massachusetts Institute of Technology (Cambridge, MA). Patient consent was not sought because the study did not impact clinical care and protected health information was de-identified in compliance with the Health Insurance Portability and Accountability Act (HIPAA). The database includes information such as demographics, vital sign measurements made at the bedside, laboratory test results, procedures, medications, and nurse and physician notes. We used the MIMIC Code Repository to query the database in PostgreSQL 10. Only those patients with a single ICU admission per hospital stay, staying more than 12 hours in the ICU, and older that 15 years old were considered for the analysis. Because the age of patients older than 89 at their first admission has been fixed to 300 as part of the de-identification process, we set their age to the median value for that group of 91.4.

We divided the dataset randomly into train, validation, and test. Given that a patient can have one or more hospitalizations with ICU stays between 2001 and 2012, we divided the dataset based on the subjects rather than on the ICU admissions. Thus, patients with more than one hospital stay remained in the same subset of data. The training sample consisted of 60% of patients, while the validation and test samples consisted of 20% patients each. We used the same dataset split across all the models presented in this article.

### b) Structured features

Our prediction algorithms included the 14 variables from the SAPS II score: age, Glasgow coma scale, systolic blood pressure, heart rate, body temperature, PaO2/FiO2 ratio, urinary output, serum urea nitrogen level, white blood cell count, serum bicarbonate level, sodium level, potassium level, bilirubin level, and type of admission (scheduled surgical, unscheduled surgical, or medical). We did not include the three underlying disease variables (acquired immunodeficiency syndrome, metastatic cancer and hematologic malignancy) because these are usually derived from the ICD9-codes, which we were predicting as well. Instead of using SAPS II variable scores, we took the worst value of those variables within the first 24 hours in the ICU, as defined by Le Gall et al. (1993). We inspected the variables for missing values and outliers. Because missing values may contain hidden patterns, we created a new dummy variable for every variable with more than 5% of missing data were 1: “missing value” and 0: “not missing”. Bilirubin, PaO2/FiO2 ratio, and urinary output had 56.1, 64.8 and 5.3 % missing values. We followed clinical knowledge to replace missing and outlier values with the mean value of the feature. Missing categorical variables were replaced with the most frequent value. All continuous variables were standardized by first obtaining the parameters from the training dataset and applying the transformation on training, validation and test.

### c) Unstructured features

We included two different unstructured features to help better represent the patients’ state upon admission to the ICU. The ADMISSIONS table in MIMIC-III contains a preliminary diagnosis for patients upon hospital admission. This field is usually assigned by the admitting clinician, does not have a systematic format, and it varies between 1 and 22 terms. We are aware that the ‘diagnosis’ field could be vague and that the documentation recommends to not use this field to stratify patients. However, we think that whether this field is useful will be determined during training. The other unstructured feature was obtained from the NOTEEVENTS table. We analyzed the structure of the discharge summaries and built a parser to extract the PAST MEDICAL HISTORY of all patients (details in our repository). Medical histories were already de-identified, and ranged from 0 to ∼720 words. We truncated the medical history feature to 242 words, which includes 99% of all notes. We used pre-trained, 200 dimension word embeddings (Zhang et al. 2019) to initialize the model after tokenizing the words in the vocabulary (see our repository for more details on the text normalization). The vocabulary was defined by the collection of terms present only in the training set. All new terms appearing in the validation and test datasets were treated as unknown and thus got the same index and embedding representation.

### d) Prediction Models

We describe two models for mortality prediction using unstructured features. Both models use a GRU structure (Cho et al. 2014) to capture important features from the text. Given a sequence of text, each pretrained word embedding passes as an input to the GRU at each timestep t. The GRU stores information in a hidden layer h that will be updated overtime. The output of the hidden layer at the final timestep, t, is the final hidden state vector ht, which is then combined with other features to estimate ŷ. The two models are very similar in the architecture of the input and hidden layers, but they differ in the classification task, as explained below. We implemented our deep learning models in PyTorch (Paszke et al. 2019). We used the training set for training different models, the validation set for hyperparameter tuning, and the test set as a held-out set. The results presented here correspond to the model evaluation metrics performed on the test set.

#### Deep learning Model for mortality prediction

This is a binary classifier that combined unstructured and structured features to predict the probability of mortality of each ICU admission (Figure 1). We concatenated the final hidden state vectors from two GRUs (*hts*) with an 18-dimension vector (14 SAPS variables + 3 dummy variables from variables with missing values), and a 11-dimension embedding representing 3 categorical variables (Gender, Admission type, ICU type and Admission location). This concatenated vector was fed into a fully connected layer, normalized, and a ReLu activation layer. Lastly, a fully-connected layer was fed to a sigmoid layer which output the probability distribution over the label. Dropout was added at different steps to reduce model overfitting. The best performing model on the validation set had the following parameters: dimension of the GRU hidden layer: 20, dimension of hidden layer in the fully connected layer: 150, batch size: 3500, dropout probability: 0.5, learning epochs: 35, learning rate: 0.005 (5 epochs), 0.01 (15 epochs), 0.005 (5 epochs), and 0.001 (10 epochs). We used a binary cross entropy loss and Adam optimizer (Kingma and Ba 2014).

**Figure 1.**
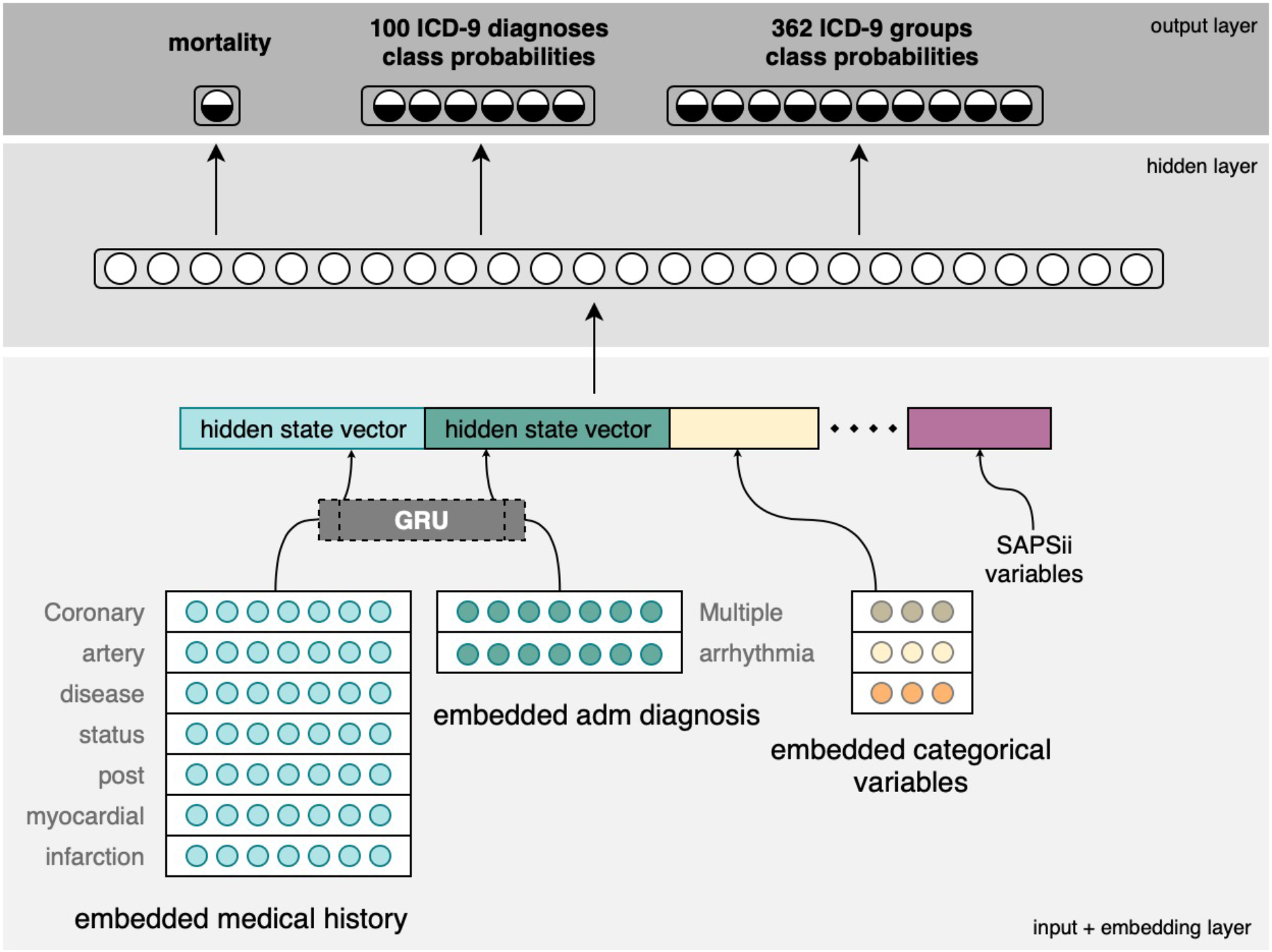
Deep Learning Model architecture for mortality prediction in Intensive Care Units. Unstructured data from medical history records (NOTEEVENTS) and admission diagnosis (ADMISSIONS) are processed by a GRU layer, then the h*t* vectors are concatenated to the tabular variables used in the SAPSII score. A fully connected hidden layer and an activation function computes the probability of mortality during the ICU stay. A multitask model simultaneously predicts mortality and ICD9 codes.

#### Deep learning Multitask model for ICD9 assignment and mortality

The input layers of this model are similar to the model described in the previous paragraph. However, the prediction from this model was a combination of 3 tasks, also known as multitask training (Figure 1). The first task was mortality and the other two were multi-label classifiers for different groups of ICD9-codes, as follows. Of the ∼13,000 existing ICD9-codes, about 6,300 distinct ones where assigned to the patients in MIMIC. The number of diagnoses per patient and ICU admission was variable, and ranged between 1 and 39 codes. This means that one or more can be assigned to any given ICU stay. Because many of these diagnostic codes were rare, we focused on predicting the most common 100 as our second task. Even with this limitation, some of those diagnoses appeared in only 3% of the training data. The remaining codes were coerced to their category group. We kept 362 of those groups as our third task, where each group had an absolute frequency of at least 100 of ICU admissions. The best performing model had the following parameters: dimension of the GRU hidden layer: 20, dimension of hidden layer in the fully connected layer: 300, batch size: 3500, dropout probability: 0.5, learning epochs: 50, learning rate: 0.005 (5 epochs), 0.01 (25 epochs), 0.005 (10 epochs), and 0.001 (10 epochs). Each task has its own binary cross entropy loss, and then the three losses were summed to obtain the total loss at each training step. We used a heuristic approach to find a linear combination of the loss function so that the losses had approximately a similar scale. This allowed the tasks to equally affect the learning process. The weights of these hyperparameters were: mortality loss: 2.5, 100 most frequent ICD9 loss: 1.3, and 362 groups ICD9 loss: 2.

### e) Baseline Evaluation and Performance Measures

A logistic regression model, an eXtreme Gradient Boosting model (XGBoost, Chen and Guestrin 2016), and a neural network model were fit to the structured data using the same training, validation, and test sets as for the neural network models. These three algorithms were compared to our proposed deep leaning models. The outcome measure of the logistic regression baseline model was hospital mortality. The traditional SAPS II uses a logistic regression on the SAPS II score with pre-defined coefficients (Le Gall, Lemeshow, and Saulnier 1993). In our version of the SAPS II logistic regression, we used the values of the variables instead of the variable’s scores. Categorical variables were one-hot encoded. We used cross-validation with 5 folds and L1 regularization to find the model hyperparameters.

We used XGBoost as a baseline for the multitask neural network models. The outcome measure of the XGBoost baseline model was hospital mortality and labels for the ICD-9 codes, but in contrast to the neural network, the XGBoost fits one classifier per class, in a one-versus-rest manner and it does not handle the problem when the same observations have multiple classes assigned (multi task learning). We then took the mean of the evaluation metric for the two ICD9 tasks to be able to compare among models. The XGBoost model had the following parameters: minimum child weight: 40, learning rate: 0.05, fraction of columns sampled by tree: 0.3, maximum tree depth: 4, and random subsample fraction: 0.8. Lastly, to account for high class imbalance, we set the *scale pos weight* to be the ratio between the negative classes and the positive classes for each classifier.

Lastly, a feed-forward neural network was trained on the structured data to predict either hospital mortality, or mortality and ICD9-codes in a multitask model architecture. In this model, we concatenated an 18-dimension vector (14 SAPS variables + 3 dummy variables from variables with missing values), and a 11-dimension embedding representing 3 categorical variables (Gender, Admission type, ICU type and Admission location). This concatenated vector was fed into a fully connected layer, normalized, and a ReLu activation layer. Lastly, a fully-connected layer was fed to a sigmoid layer which output the probability distribution over the label. The best performing model for hospital mortality alone had the following parameters: dimension of hidden layer in the fully connected layer: 150, batch size: 3500, dropout probability: 0.5, learning epochs: 30, learning rate: 0.005 (5 epochs), 0.01 (15 epochs), 0.005 (5 epochs), and 0.001 (10 epochs). The multitask learning model had the following parameters: dimension of hidden layer in the fully connected layer: 150, batch size: 3500, dropout probability: 0.5, learning epochs: 60, learning rate: 0.01 (30 epochs), and 0.005 (30 epochs).

A good model with high discrimination power will assign a higher probability to a patient that died in the hospital, or a higher probability to an ICD9-code that was assigned at discharge. The discrimination power of the algorithms was assessed with the area under the receiver-operating characteristic curve (AUROC), reported with 95% CI. The 95% CI were obtained by bootstrapping the test dataset with replacement 100 times. Another model performance of interest is calibration. Under perfect calibration, there is a match between the average predicted probability and the observed probability at each probability range. Model calibration was only calculated for the deep learning model with structured and unstructured data.

### f) Patient phenotyping evaluation

Machine learning models with high interpretability are highly desired in clinical informatics. In the context of this research, we want to evaluate whether the GRU was able to extract relevant information about patients’ clinical phenotypes for each hospital admission. In other words, has the algorithm learned something useful? If the algorithm were able to differentiate patients’ clinical phenotypes, this would suggest that any improvement in model predictions was due to a better segmentation of patients’ ICU stays into risk groups. For this evaluation, we used a different set of hospital notes from the discharge summary of the NOTEEVENTS table to group patients stays according to their clinical phenotypes (see Table 1 for an example). We parsed the discharge summaries, searched, and extracted the following sub-sections for each patient: a) discharge condition, b) discharge diagnosis, c) final diagnosis, and d) history of present illness. These sub-sections made up a separate dataset that has not been previously seen by the model to make predictions. The groupings generated with this dataset were compared to groupings derived from the GRU model ht vector.

**Table 1.** Examples of the unstructured features used in this study. Past Medical History was used as input in the GRU-Neural Network, and the Discharge Diagnosis was used to group patients in risks groups with LDA modeling.

|  | Past Medical History | Discharge Diagnosis |
| --- | --- | --- |
| Patient A | 1. Interstitial pulmonary fibrosis.<br>2. T7 compression fracture.<br>3. No cardiac history. | “The patient is a 54-year-old man with a history of interstitial pulmonary fibrosis, UIP who presents with fever and sudden shortness of breath. The patient has shortness of breath at baseline, requiring home oxygen of two to four liters. He is on chronic prednisone treatment, gamma interferon for his interstitial pulmonary fibrosis...()” |
| Patient B | Dyslipidemia with triglycerides in dm type II macular degeneration htn kidney stone gastritis cervical spondylosis colonic polyp | “Stable. Primary: brain tumor. Secondary: dyslipidemia tg type II dm macular degeneration htn kidney stone gastritis cervical spondylosis colonic polyp gout 62 yo male w/ pmhx sig for dm ii, htn, and severe hyperlipidemia who was found down at home. Pt was apparently last seen well by his friends on Saturday. When he did not show up for a golfing outing with friends, they went to his house and found him seizing behind the house on a bed of trash.” |

To make the two groupings, we first clustered patients stays into broadly-defined risks groups with topic modeling using the available post-admission information, that is, the sub-sections mentioned above (LDA, Blei, Ng, and Jordan 2003). Second, we independently grouped patients stays by applying K-means classification to the ht vectors from the GRU model, which were learned from the patient’s past medical history (see Unstructured Features section). We generated different cluster scenarios by changing the number of K-mean clusters. Third, we used the normalized mutual information score (NMI) to assess the similarity of the K-means clustering to the LDA topic assignment. A NMI score of zero means that there is no agreement between the two groupings, while a NMI score of 1 implies that there is a perfect correlation.

## 3) Results

### ICU stays

This study included 42,601 ICU stays, corresponding to 33,973 different patients. Median age was 65 (IQR = 52-78) and male patients were the majority (70%). The median SAPS II at admission was 33 (24-42). Patients were hospitalized mostly at the MICU: CCU (5809, 13.6%), CSRU (7663, 18%), MICU (17404, 40.8%), SICU (6847, 16.1%), and TSICU (4878, 11.5%). The number of deaths in ICU was 4,190, an estimated hospital mortality of 9.8%.

### Baseline and Prediction Models evaluation

The AUROC from hospital mortality prediction was 0.85 for the Logistic Regression, and 0.88 for the XGboost model (Table 2). Our baseline model achieved similar results compared to the predictions obtained by the Super Learner, Pirracchio et al. (2015). There were no significant differences between the XGboost AUROC and the AUROC obtained with a Neural Network model using the same tabular data as input variables. Moreover, we did not find any AUROC improvement when mortality was simultaneously trained with ICD9 codes compared to the XGboost one-versus-rest (Table 1, Neural Network tabular Multitask Learning vs. XGboost. The Neural Network model which included the medical history outperformed all the other models (AUROC = 0.90, Table 1). Lastly, the inclusion of medical history to the multitask learning led to a greater improvement of the ICD9 codes prediction than of the mortality prediction (Multitask Learning, ICD task vs. Mortality task trained with tabular data only or tabular + unstructured data, Table 2).

**Table 2.**
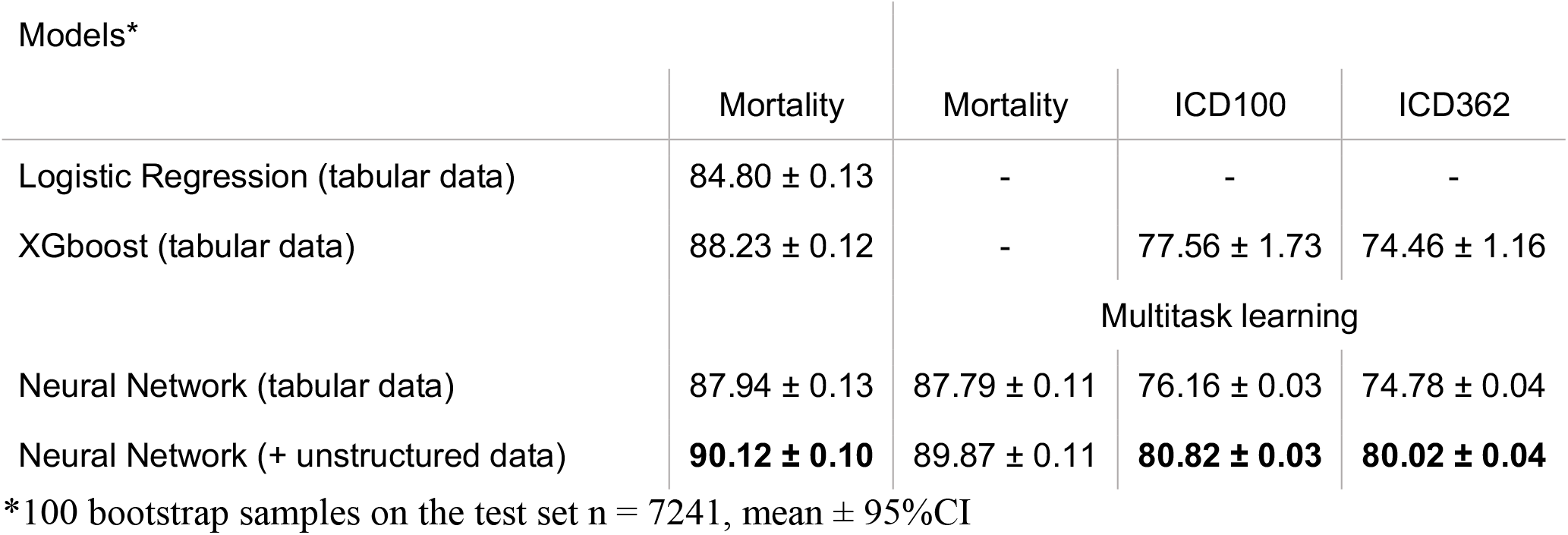
Classification performance (% AUCROC) of prediction models. Logistic Regression and XGboost models were fitted with tabular data only. Neural Network models were fitted with and without patients’ medical history and compared to the baseline models.

The calibration plot shows that the Deep learning Multitask model provides accurate predictions throughout the range of death probability (Figure 2). The predicted probabilities fell close to the ideal calibration line for low probabilities of death, and were slightly underestimated as probabilities increased.

**Figure 2.**
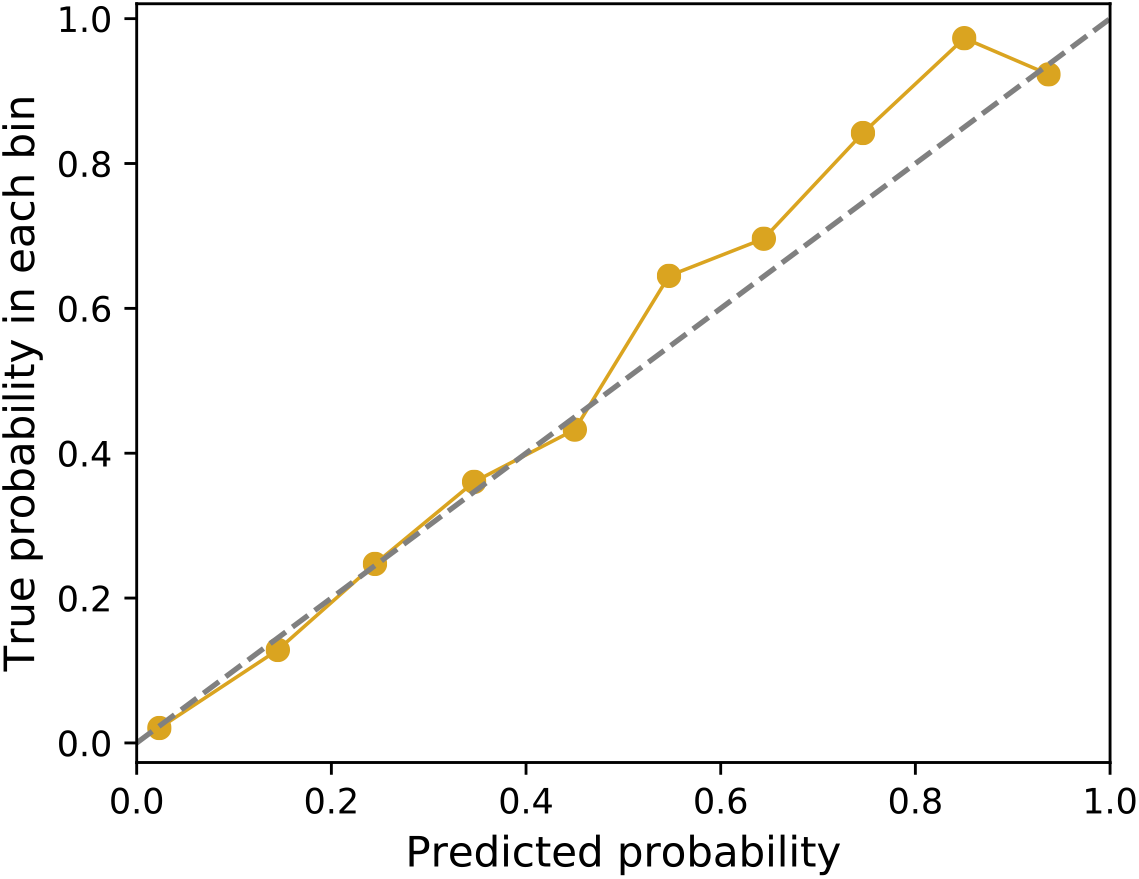
Calibration plot for the actual death probability compared the mortality prediction given by the Deep learning Multitask model.

### GRU Model Interpretability and Patient Phenotyping

To improve understanding of the factors contributing to the prediction in our model, a set of notes with the patients’ condition and diagnosis at discharge was used to cluster the patients into risk groups. We built several LDA models with the number of topics ranging from 8 to 20. Although the model with 14 topics had the highest coherence scores (∼0.55), the keywords describing these topics did not create conceptually distinct risks groups compared to a model with 12 topics with a coherence score of 0.54 (Figure 3). The mean NMI score between the LDA topics and the k-means clustering was 0.24 (Figure 4). The NMI score for the LDA topics and a randomly assigned cluster number was not different from zero. From the 12 topics, we could identify a subset of 7 groups related to the following conditions: coronary artery disease, sepsis, cancer, liver disease, respiratory problems, falls + fractures, and renal issues. When we subset the ICU stays to these 7 groups, the mean NMI core was ∼0.38 (Figure 4). The positive correlation between the clusters based on patients’ discharged summaries and the clusters based on past medical history shows that the GRU based clusters are effective at segmenting patients based on past medical history.

**Figure 3.**
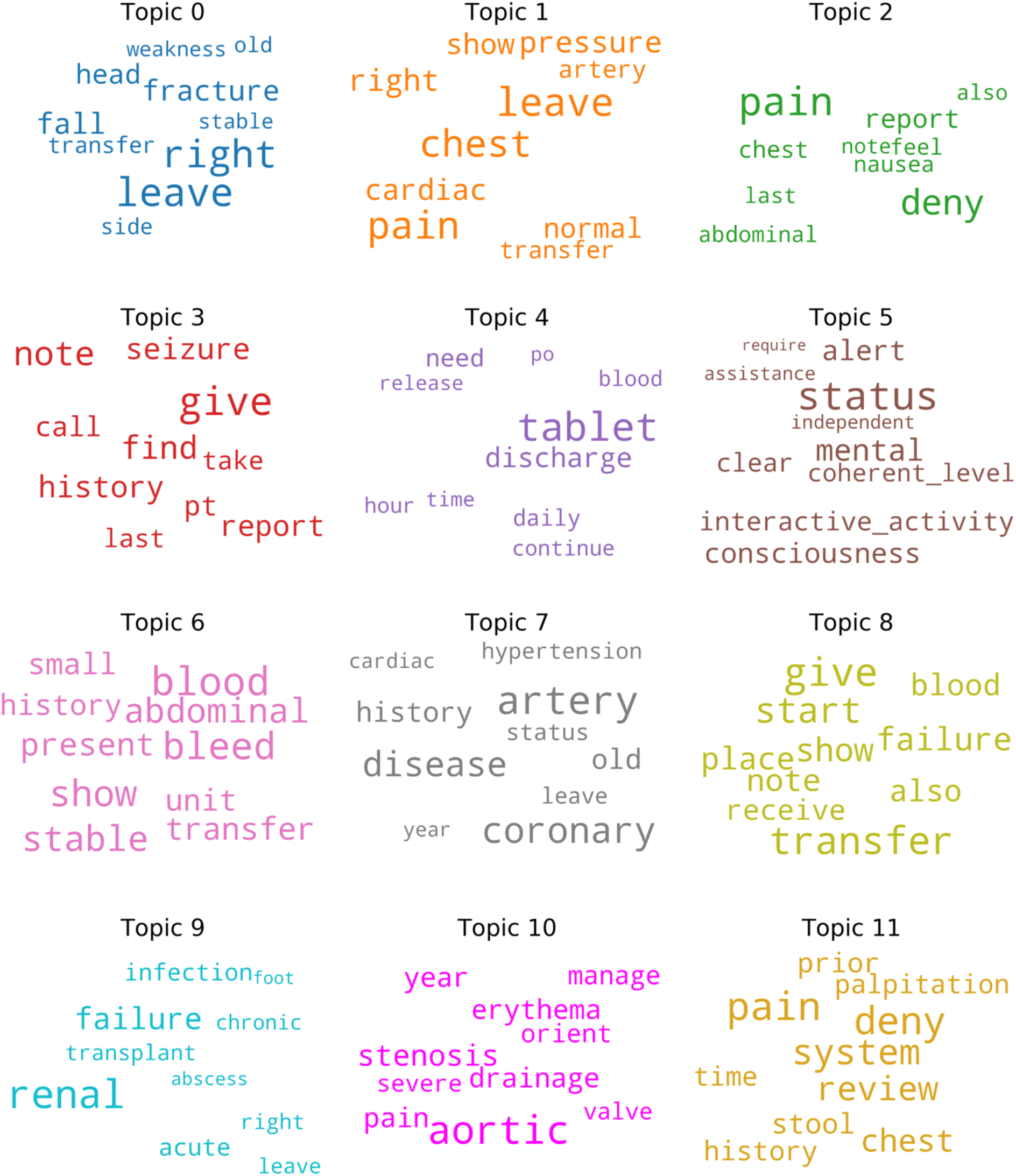
Visualization of the topics generated with LDA modeling. For each of the 12 topics, a word cloud shows the 10 top keywords where the size of the words is proportional to their weight in the topic. Some topics can be associated with risks groups The topics Topic visualization. coronary artery disease, sepsis, cancer, liver disease, respiratory problems, falls + fractures, and renal issues

**Figure 4.**
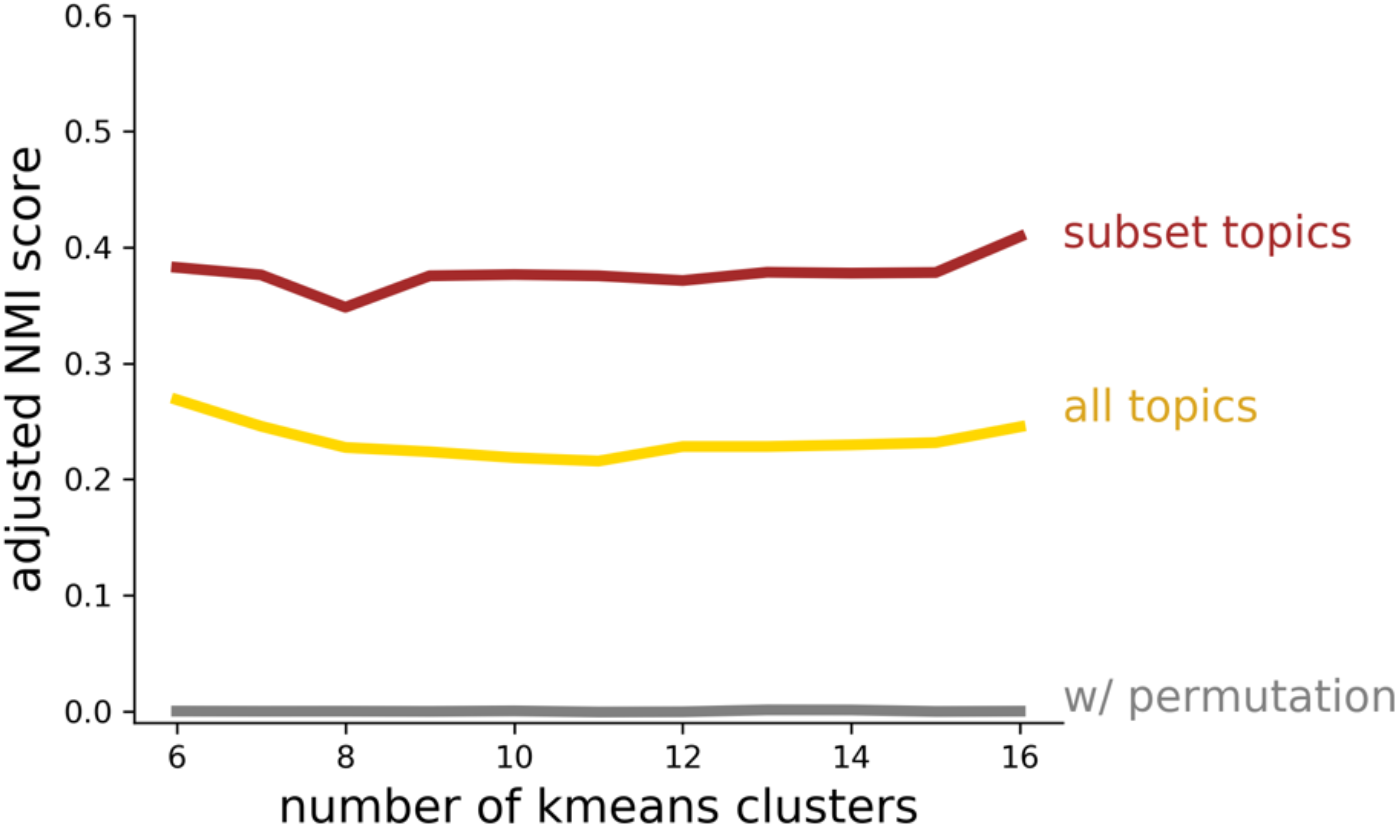
The adjusted normalized mutual information score (NMI) for various K-means clusters grouping the ht vectors derived from patients’ medical histories and the LDA topics obtained by analyzing the patients’ discharge summaries. In yellow, predicted topics on the test set ICU stays were compared to clusters obtained with K-means after computing the ht vectors. In red, the same K-mean clusters were compared to a subset of 7 predicted topics (likely corresponding with: coronary artery disease, sepsis, cancer, liver disease, respiratory problems, falls + fractures, and renal issues). In grey, the adjusted NMI was calculated by randomly permutating the K-means cluster assignment, we expect low correlation between these.

## 4) Discussion

We have assessed the potential of medical history data to improve outcome predictions of critically ill patients in the ICU. Our models with tabular data as the only input variables achieved similar results to those previously published (Pirracchio et al. 2015, Le Gall, Lemeshow, and Saulnier 1993, Sakr et al. 2008). As severity scores need to account for, and be updated to, common comorbidities, we hypothesized that past medical history would allow a simple way to include comorbidity information, account for cumulative impacts, or adjust for different patients’ underlying risks. Our experiments showed that the inclusion of the medical history resulted in significant higher AUCROC for in-hospital mortality prediction. Moreover, this model which uses medical history notes and 24h of ICU physiological variables, showed similar mortality prediction as models that included 48 hours of ICU’s notes (Weissman et al. 2018). The advantage of using recurrent neural networks such GRUs, is that GRUs can process inputs of any length, and without much pre-processing work.

Our results did not support the hypothesis that including diagnosis codes as outcomes would improve mortality predictions by means of multitask learning. While including the medical history significantly improved the automated diagnosis coding, multitask learning did not have an effect on improving the main outcome of interest, prediction of mortality. Interestingly, the relative improvement in diagnosis coding due to including patients’ medical history was greater than the improvement in mortality. This results supports the hypothesis that most of the patients’ note are included to justify the billing and as such they correlate more with ICD codes than with mortality. This will indicate that for Machine Learning to be able to harness the true potential of our Electronic Health Record a pattern shift of how physicians input patients’ initial information would be needed. Needless to say, this is a major undertaking.

Our findings not only supported the use of unstructured notes to improve outcome predictions, but also provided an interpretation of the mechanism. We think that the inclusion of medical history processed by the GRU memory mechanism results in a model customized for different risks groups as also reported by other authors (Lee and Maslove 2017). This is supported by the correlation found between patients that were grouped according to discharge diagnoses with the grouping of patients produced by the GRU.

## Data Availability

We used publically available data from MIMIC dataset.

